# Modeling and Prediction of the 2019 Coronavirus Disease Spreading in China Incorporating Human Migration Data

**DOI:** 10.1101/2020.02.18.20024570

**Authors:** Choujun Zhan, Chi K. Tse, Yuxia Fu, Zhikang Lai, Haijun Zhang

## Abstract

This study integrates the daily intercity migration data with the classic Susceptible-Exposed-Infected-Removed (SEIR) model to construct a new model suitable for describing the dynamics of epidemic spreading of Coronavirus Disease 2019 (COVID-19) in China. Daily intercity migration data for 367 cities in China are collected from Baidu Migration, a mobile-app based human migration tracking data system. Historical data of infected, recovered and death cases from official source are used for model fitting. The set of model parameters obtained from best data fitting using a constrained nonlinear optimization procedure is used for estimation of the dynamics of epidemic spreading in the coming weeks. Our results show that the number of infections in most cities in China will peak between mid February to early March 2020, with about 0.8%, less than 0.1% and less than 0.01% of the population eventually infected in Wuhan, Hubei Province and the rest of China, respectively.

## 1. Introduction

The Coronavirus Disease 2019 (COVID-19) (known earlier as New Coronavirus Infected Pneumonia) began to spread since December 2019 from Wuhan, which has been widely regarded as the epicenter of the epidemic, to almost all provinces throughout China and 28 other countries. Human-to-human transmission had been found to occur in some early Wuhan cases in mid December [1], and the high volume and frequency of movement of people from Wuhan to other cities and between cities is thus an obvious cause for the wide and rapid spread of the disease throughout the country. The Susceptible-Exposed-Infected-Removed (SEIR) model has traditionally been used to study epidemic spreading with various forms of networks of transmission which define the contact topology [2], such as scalefree networks [3, 4, 5], small-world networks [6, 7], Oregon graph [8, 9], and adaptive networks [10]. Moreover, in most studies, the contact process assumes that the contagion expands at a certain rate from an infected individual to his/her neighbor, and that the spreading process takes place in a single population (network). The COVID-19 outbreak, however, began to occur and escalate in a special holiday period in China (about 20 days surrounding the Lunar New Year), during which a huge volume of intercity travel took place, resulting in outbreaks in multiple regions connected by an active transportation network. Thus, in order to understand the COVID-19 spreading process in China, it is essential to examine the human migration dynamics, especially between the epicenter Wuhan and other Chinese cities. A recent study has also revealed the risk of transmission of the virus from Wuhan to other cities [11].

In this paper, we utilize the human migration data collected from Baidu Migration [12], which provides historical indicative daily volume of travellers to/from and between 367 cities in China. To demonstrate the impact of intercity traffic on the COVID-19 epidemic spreading, we plot in Figure 1 the number of infected individuals in different cities versus the inflow traffic volume from Wuhan, which clearly shows that for cities farther away from Wuhan, the number of infected individuals almost increases linearly with the inflow traffic from Wuhan. In view of the importance of human migration dynamics to the disease spreading process, we combine, in this study, intercity travel data collected from Baidu Migration [12] with the traditional SEIR model [2] to build a new dynamic model for the spreading of COVID-19 in China. Using official historical data of infected, recovered and death cases in 367 cities, we perform fitting of the data to estimate the best set of model parameters, which are then used to estimate the number of individuals exposed to the virus in each city and to predict the extent of spreading in the coming months. Our study shows that the number of infected cases in various Chinese cities will peak between mid February to early March 2020, with about 0.8%, less than 0.1% and less than 0.01% of the population eventually infected in Wuhan, Hubei Province, and the rest of China, respectively.

**Figure 1:**
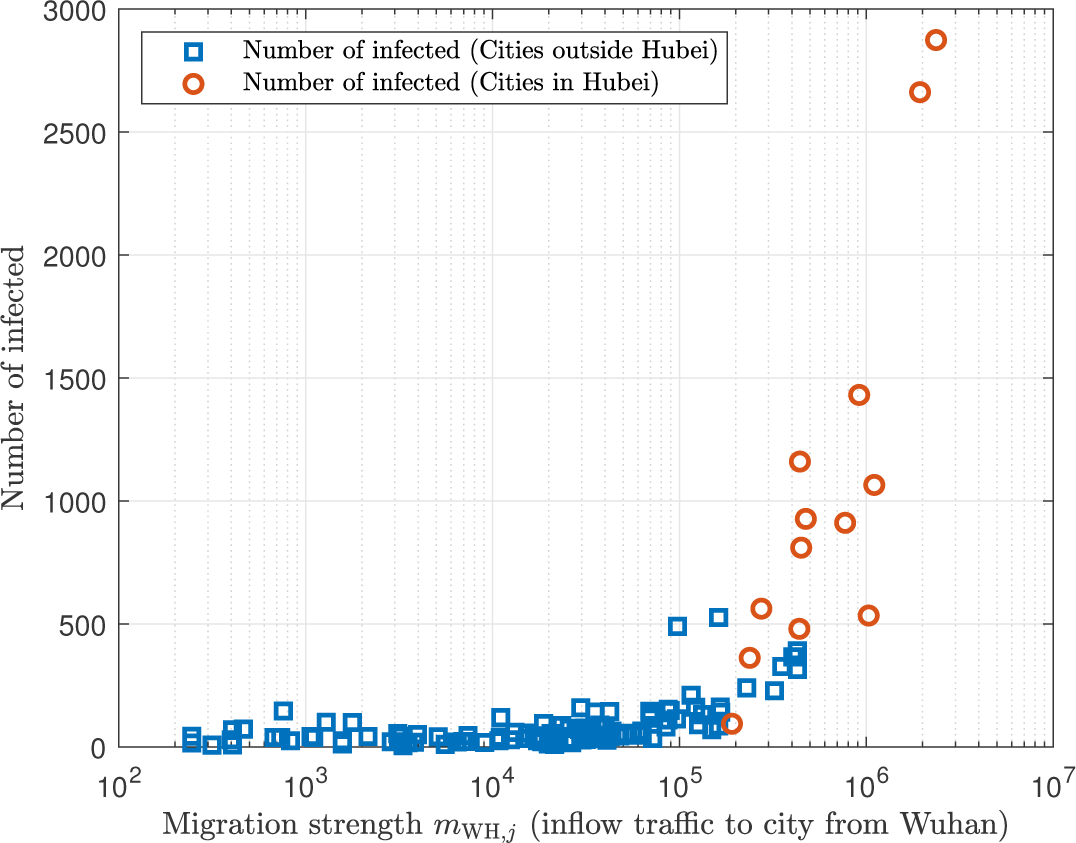
Number of infected individuals in various cities on 13/Feb/2020 versus the city’s inflow traffic from Wuhan. Inflow traffic of each city from Wuhan is quantified by migration strength from Wuhan extracted from Baidu Migration data.

In the remainder of the paper, we first introduce the official daily infection data and the intercity migration data used in this study. The SEIR model is modified to incorporate the human migration dynamics, giving a realistic model suitable for studying the COVID-19 epidemic spreading dynamics. Historical data of infected, recovered and death cases from official source and data of daily intercity traffic (number of travellers between cities) extracted from Baidu Migration are used to generate the model parameters, which then enable estimation of the propagation of the epidemic in the coming months. We will conclude with a brief discussion of our estimation of the propagation and the reasonableness of our estimation in view of the measures taken by the Chinese authorities in controlling the spreading of this new disease.

## 2. Data

### 2.1. Official Data of COVID-19 Cases

The availability of official data of infected cases in China varies from city to city. Wuhan, being the epicenter, has the first confirmed case of COVID-19 infection on December 8, 2019 [1]. Most other cities in China began to report cases of COVID-19 infections around mid January 2020. Our data of daily infected and recovered cases, and death tolls, are based on the official data released by the National Health Commission of China, and the daily data used in our study are from January 24, 2020, to February 16, 2020, including the daily total number of confirmed cases in each city, daily total cumulative number of confirmed cases in each city, daily cumulative number of recovered cases in each city, and daily cumulative death toll in each city. It should be emphasized that the official data may not be the actual (true) data. Although the earliest confirmed case appeared on December 8, 2019, subsequent missing cases were expected to be significant in Hubei Province in the early stage of the epidemic outbreak. Systematic updates of infection data in other cities began after January 17, 2020. Figure 2 shows the number of confirmed infected cases, recovered cases and death tolls of six major Chinese cities.

**Figure 2:**
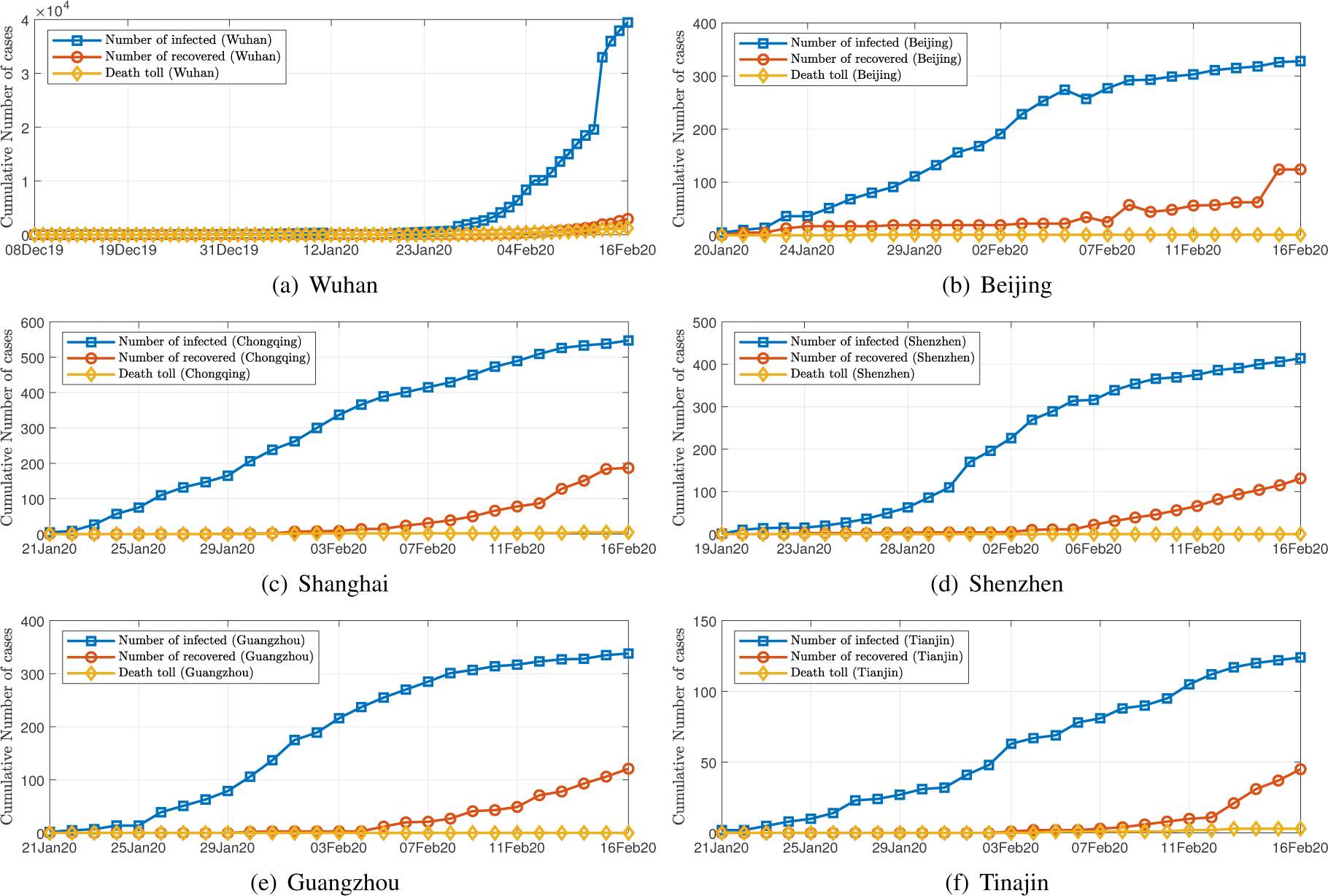
Daily data of COVID-19 infections in six Chinese cities from December 8, 2019 to February 13, 2020. (a) Wuhan (available from December 8, 2019); (b) Beijing (available from January 20, 2020); (c) Chongqing (available from January 20, 2020); (d) Shenzhen (available from January 19, 2020); (e) Guangzhou (available from January 21, 2020); (d) Tianjin (available from January 21, 2020).

### 2.2. Intercity Travel Data

As human-to-human transmission has been confirmed to occur in the spreading of COVID-19, gatherings of people and intercity travel of infected and exposed individuals within China have been the main drives that escalate the spreading of the virus. The period (around 20 days) surrounding the Lunar New Year (mid January to early February in 2020) is the most important holiday period in China. Migrant workers and students travel from major cities to country towns for family reunions, and return to the cities at the end of the holiday period. Holiday goers also travel to and from tourist cities. China’s Ministry of Transport estimates around 3 billion trips to be taken during this period. Wuhan, being a major transport hub and having a large number of higher education institutions as well as manufacturing plants, is among the cities with the largest outflow and inflow traffic before and after the Chinese New Year festival. Our study aims to incorporate these important human migration dynamics in the construction of the spreading model. We collect daily intercity travel data in China from Baidu Migration, which is a mobile-app based big data system recording movements of mobile phone users. Specifically, we have collected Baidu Migration data for 367 cities (or administrative regions) in China over the period of January 1, 2020, to February 13, 2020. The data provide the migration strengths of cities which are indicative measures of the human traffic volume moving in and out of individual cities and administrative regions, as depicted by the inflow and outflow networks shown in Figure 3(a).

**Figure 3:**
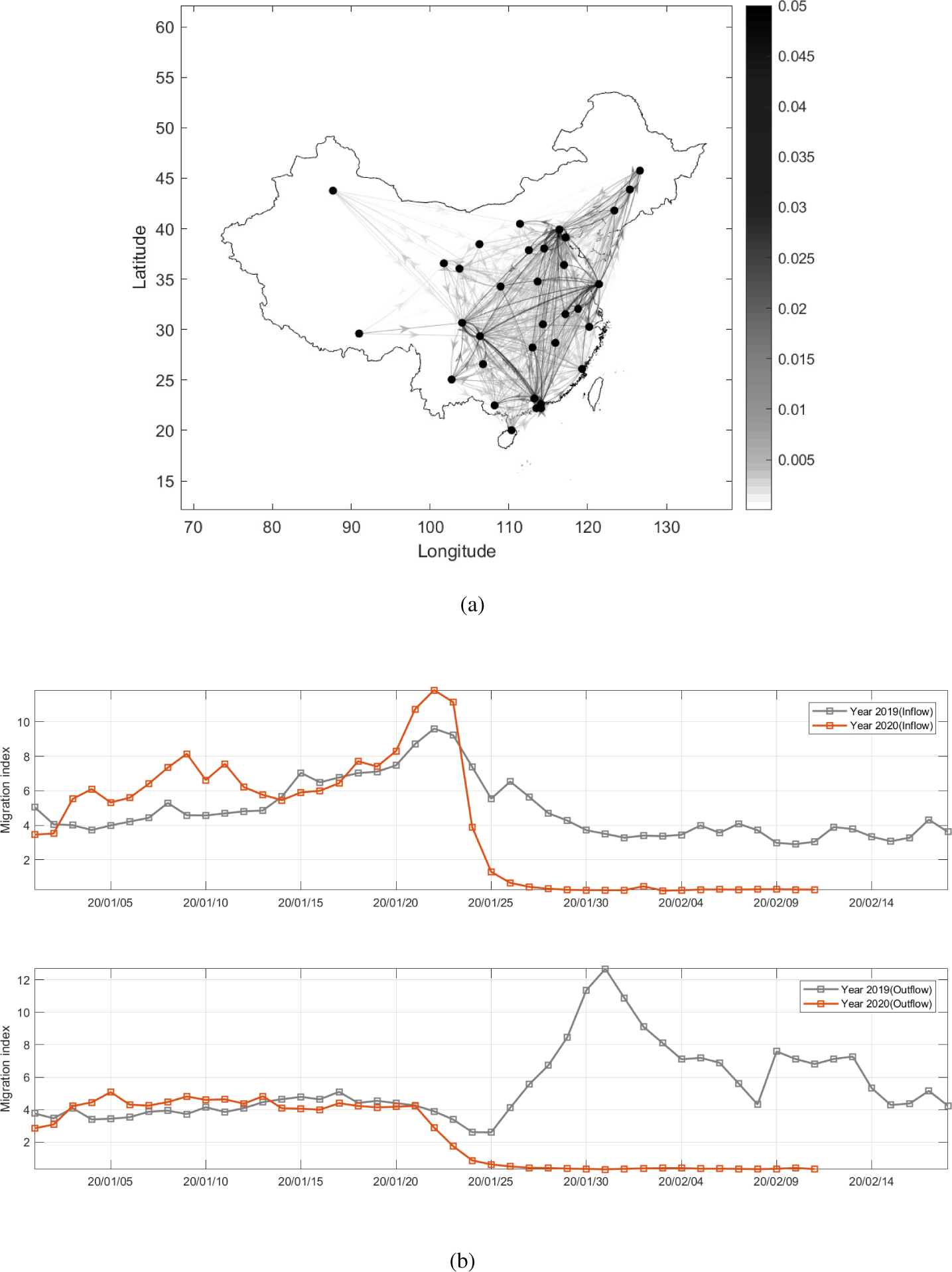
(a) Illustration of travel network of major cities in China with arrows indicating direction of travel and darkness of lines indicating migration strengths (relative traffic volumes); (b) total inflow/outflow of travellers to/from Wuhan from/to other Chinese cities using Baidu Migration data. Inflow and outflow data of each city with individual cities are also collected to form *m*_*i j*_.

Based on the collected data, we construct the *migration matrix*, which is given as

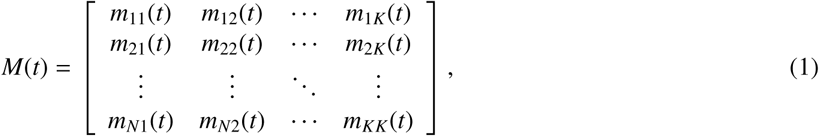

where *K* is the number of the cities or administrative regions (*K* = 367 in this study), and *m*_*i j*_(*t*) is the migrant volume from city *i* to city *j* at time *t*. Migration matrix *M* thus effectively describes the network of cities with human movement constituting the links of the network, as shown in Figure 3(a). Several properties of *M* are worth noting:

- *M* records migration from one city to another. Movement within a city is not counted, i.e., *m*_*ii*_(*t*) = 0 for all *i*.
- *M* is non-symmetric as traffic from one city to another is not necessarily reciprocal at any given time, i.e., *m*_*i j*_(*t*) ≠ *m*_*ji*_(*t*).
- Number of outflow migrants of city *i* at time *t* is

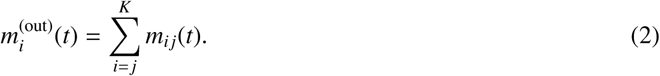

- Number of inflow migrants of city *i* at time *t* is

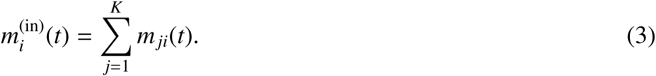

The charts in Figure 3(b) plot the daily total inflow and outflow migration strengths of Wuhan, showing the abrupt decrease of migration strengths after the city shut down all inbound and outbound traffic from January 24, 2020. A visualization of migration matrix *M* is given in Figure 4, where x-axis and y-axis correspond to city *i* and city *j*, respectively, and the color intensity indicates the migrant volume.

**Figure 4:**
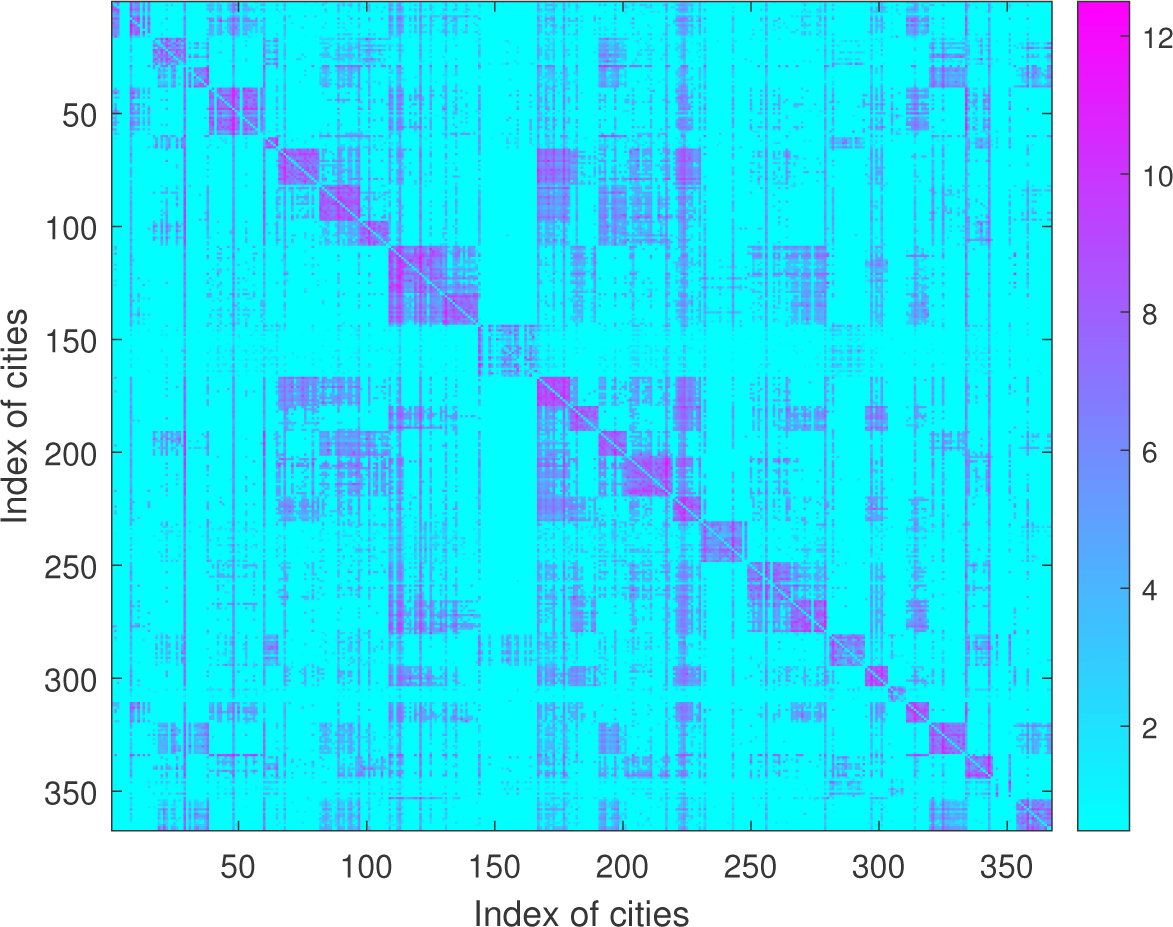
Visualization of migration volume (log(*m*_*ij*_)) from city *i* (x-axis) to city *j* (y-axis) on January 23, 2020.

## 3. Method

### 3.1. Model

In the SEIR model, each individual in a population may assume one of four possible states at any time in the dynamic process of epidemic spreading, namely, susceptible (S), exposed (E), infected (I) and recovered/removed (R). The dynamics of the epidemic can be described by the following set of equations: 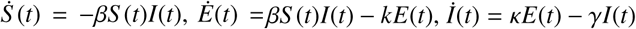 and 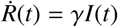, where *S* (*t*), *E*(*t*), *I*(*t*) and *R*(*t*) are, respectively, the number of people **s**usceptible to the disease, **e**xposed (being able to infect others but having no symptoms), **i**nfected (diagnosed as confirmed cases), and **r**ecovered (including death cases); *β* is the exposition rate (infection rate of susceptible individuals); *κ* is the infection rate of exposed individuals; and *γ* is the recovery rate. For simplicity, recovered individuals include patients recovered from the disease and death tolls. In discrete form, the SEIR model can be represented by

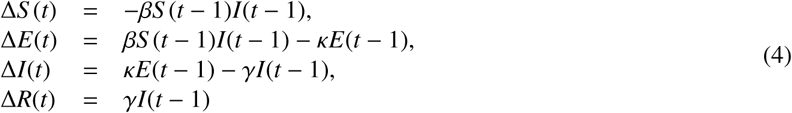

where Δ*S* (*t*) = *S* (*t*) − *S* (*t* − 1), Δ*E*(*t*) = *E*(*t*) − *E*(*t* − 1), Δ*I*(*t*) = *I*(*t*) − *I*(*t* − 1), and Δ*R*(*t*) = *R*(*t*) − *R*(*t* − 1), with *t* being a daily count. As the incubation period for COVID-19 can be up to 14 days, the number of exposed individuals (who show no symptom but are able to infect others) plays a crucial role in the spreading of the disease. The state *E*, which is not available from the official data, is thus an important state in our model. Furthermore, combining death toll with the recovered number as state *R* will simplify the computation without affecting the accuracy of our data fitting and subsequent estimation.

Suppose, for city *i*, the four states are *S* _*i*_(*t*), *E*_*i*_(*t*), *I*_*i*_(*t*) and *R*_*i*_(*t*), at time *t*. Here, we also define a total susceptible population, 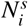, which is the eventual number of infected individuals in city *i*. Moreover, if city *i* has a population of *p*_*i*_ and the eventual percentage of infection is *δ*_*i*_, then 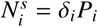. Thus, we have

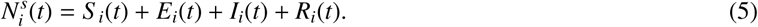

The classic SEIR model would give Δ*I*_*i*_ as the difference between the number of exposed individuals who become infected and the number of removed individuals. However, the onset of the COVID-19 epidemic has occurred in a special period of time in China, during which a huge migration traffic is being carried among cities, leading to a highly rapid transmission of the disease throughout the country. In view of this special migration factor, the SEIR model should incorporate the human migration dynamics in order to capture the essential features of the dynamics of the spreading. In particular, for city *i*, in addition to the abovementioned classic interpretation, the daily increase in the number of infected cases should also include the inflow of infected individuals from other cities, less the outflow of removed cases from city *i*. In reality, inflow and outflow of exposed individuals to and from the city are also important and to be estimated in the model. Thus, if *m*_*i j*_(*t*) people move from city *i* to city *j* on day *t*, and the population of city *i* is *P*_*i*_(*t*), then the number of infected individuals moving from city *i* to city *j* is

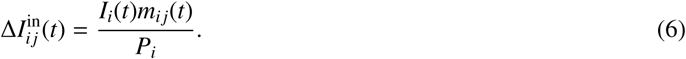

Also, the number of migrants leaving from city *j* is 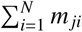, and the number of infected cases that have migrated out of city *j* is

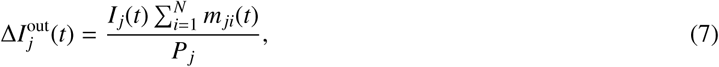

where *P*_*j*_(*t*) is the population of city *j* on day *t*. Thus, the increase in infected cases on day *t* in city *j* is given by

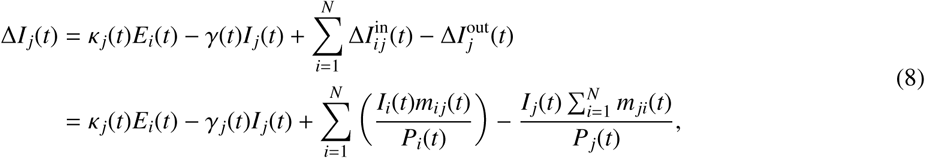

where Δ*I*_*j*_(*t*) = *I* _*j*_(*t* + 1) − *I* _*j*_(*t*) and *κ* _*j*_(*t*) is the infection rate in city *j* on day *t*, i.e., the rate at which exposed individuals become infected. Moreover, infected individuals, once confirmed, would unlikely be able to migrate to another city.

We thus implement this condition by writing (8) as

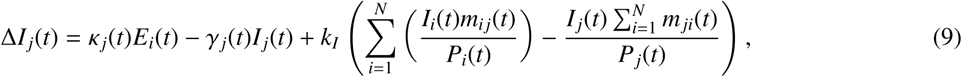

where 0 < *k*_*I*_ *≪* 1 is a constant representing the possibility of an infected individual moving from one city to another. Likewise, incorporating the migrant dynamics, the increase in exposed individuals on day *t* in city *j* is

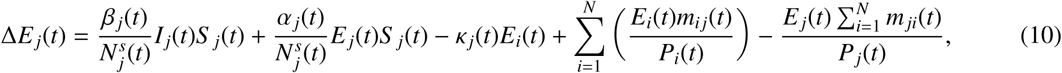

where Δ*E*_*j*_(*t*) = *E* _*j*_(*t* + 1) − *E* _*j*_(*t*), *β* _*j*_ is the infection rate of susceptible individuals in city *j*, and *α* _*j*_ is the infection rate of exposed individuals in city *j*. In a likewise fashion, we have

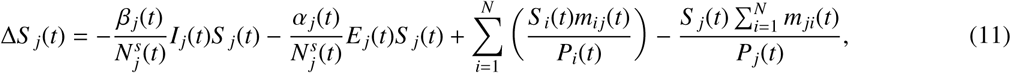

where Δ*S* _*j*_(*t*) = *S* _*j*_(*t* + 1) − *S* _*j*_(*t*). Finally, we have

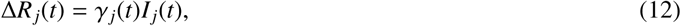

where Δ*R*_*j*_(*t*) = *R*_*j*_(*t* + 1) − *R*_*j*_(*t*). In the above derivation, we should note that

- the recovered individuals are assumed to stay in city *j*;
- the recovery rates in different cities are assumed to be different due to varied quality of treatments and availability of medical facilities;
- the recovery rates increase as time goes, as treatment methods are expected to improve gradually (i.e., taking *γ* _*j*_(*t*) as a monotonically increasing function);
- the evetual recovery rates in all cities will converge to the same constant Γ ≈ 1.

In addition, due to intercity migration, the population of city *j* on day *t* would increase or decrease according to

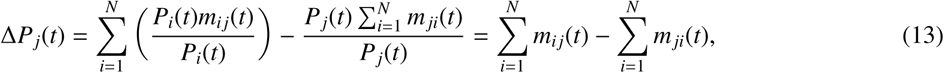

where Δ*P*_*j*_(*t*) = *P*_*j*_(*t* + 1) − *P*_*j*_(*t*). Thus, the total susceptible population should be

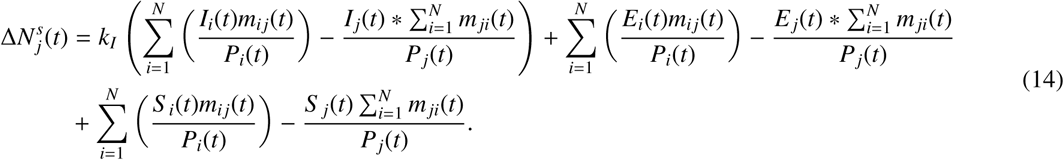

where 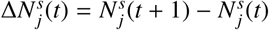.

In summary, our modified SEIR model with consideration of human migration dynamics, for city *j*, is given by

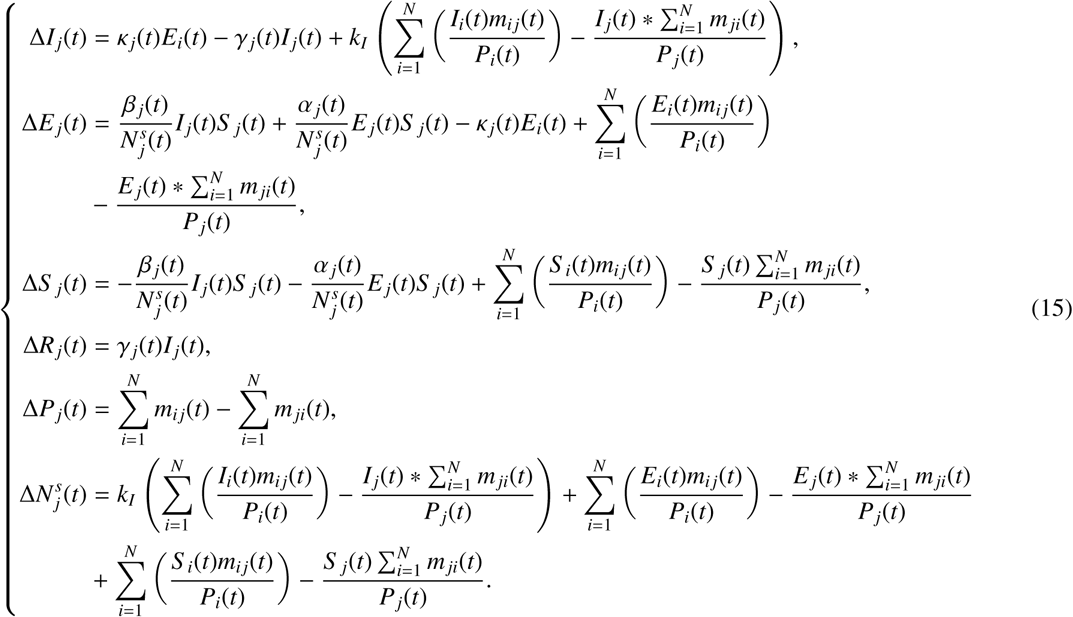

where subscript *j* denotes the city itself, and subscript *i* denotes another city from/to which people migrate on day *t*. Letting *X*_*j*_(*t*) be the extended state vector, i.e., 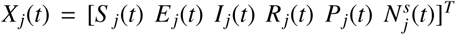, we write the above difference equation as

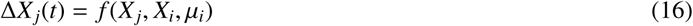

where *f* (*x*) is the right side of (15), and µ _*j*_ is the set of parameters including *α* _*j*_, *β* _*j*_, *γ* _*j*_, *κ* _*j*_ and *δ* _*j*_. For computational convenience, we write (15) as

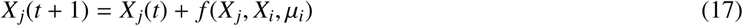

In performing the data fitting, we assume *α* _*j*_(*t*), *β* _*j*_(*t*), *γ* _*j*_(*t*), *κ* _*j*_(*t*), and *δ* _*j*_ are constants throughout the period of spreading, and the spreading begins at *t*_0_, at which 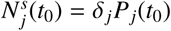.

### 3.2. Parameter Identification

The model represented by (17) describes the dynamics of the epidemic propagation with consideration of human migration dynamics. The parameters in model (17) are unknown and to be estimated from historical data. We solve this parameter identification problem via constrained nonlinear programming (CNLP), with the objective of finding an estimated growth trajectory that fits the data. An estimated number of infected cases of each city can be generated from (15) with unknown set θ _*j*_, i.e.,

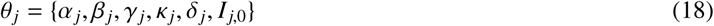

where *I* _*j*,0_ = *I* _*j*_(*t*_0_) is the initial number of infections in city *j*, and {*α* _*j*_, *β* _*j*_, *γ* _*j*_, *κ* _*j*_, *δ* _*j*_} are parameters that determine the rates of spreading and recovery in city *j*. Then, the unknown set is Θ = {*θ*_1_, *θ*_2_,…, *θ_K_*} essentially has 5*K* unknowns, where *K* is the number of cities, thus requiring an enormous effort of computation. Here, to gain computational efficiency, we assume that

- all cities share one parameter set *θ* = {*α, β, κ, γ*};
- the numbers of initial infected and exposed individuals in city *i* are *λ*_*I*_ *I*_*i*_(*t*_0_) and *λ*_*E*_ *I*_*i*_(*t*_0_), respectively, where λ_*I*_ and *λ*_*E*_ are constant;
- each city has an independent *δ*_*i*_.

Then, the size of the unknown set becomes computationally manageable, i.e.,

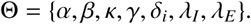

Finally, the parameter estimation problem can be formulated as the following constrained nonlinear optimization problem:

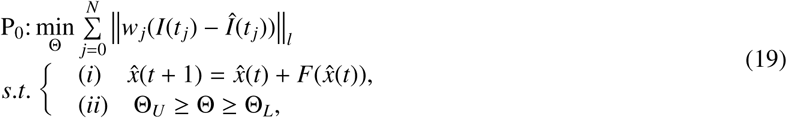

where *F*(·) represents model (15) and 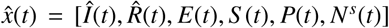 is the set of estimated variables, with unknown set Θ, which is bounded between Θ_*L*_ and Θ_*U*_. In this work, an inverse approach is taken to find the unknown parameters and states by solving (19).

The Root Mean Square Percentage Error (RMSPE) is adopted as the criterion, i.e., fitting error, to measure the difference between the number of infected individuals generated by the model and the official daily infection data.

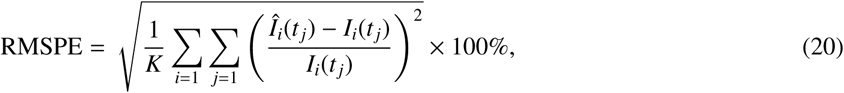

where *K* is the number of cities to be evaluated.

## 4. Results and Discussions

We perform data fitting of the model, described by (17), using historical daily infection data provided by the National Health Commission of China, from January 24, 2020 to February 13, 2020. Our approach, as described in the previous section, is to apply constrained nonlinear programming to find the best set of estimates for the unknown parameters and states. Data fitting for all 367 cities are performed. Values are updated iteratively in the optimization process. Moreover, since all parameters, like infection rates, are to be generated by fitting data with the model, the integrity of the data becomes crucial. As the official Wuhan data are expected to deviate from the true values quite significantly during the early outbreak stage due to uncertainty in diagnosis and other issues related to reporting of the epidemic by the local government, we have allowed the fitting errors for Wuhan to expand over a reasonable range, while the fitting errors for most other cities remain small. In addition, as the epidemic propagates in time, effective control measures and improved public education would reduce the infection rates for the susceptible and exposed individuals, making these parameters time varying in reality. Nonetheless, our fitting assumes these parameters being constant during the short fitting period for computational simplicity.

The propagation profiles, in terms of the number of infected individuals and estimated number of exposed individuals, for all 367 cities are estimated. As limited by space, we only show in Figure 5 the results for 20 selected cities. This model can also provide projections of the number of infected and exposed individuals in the next 200 days, as shown in Figure 6, which clearly show that the daily infection would reach a peak sooner or later. By running the identification algorithm, we identify the optimal parameter set as *α* = 0.5869, *β* = 0.8949, *κ* = 0.1008, and *γ* = 0.0602. From the estimated propagation profiles of the COVID-19 epidemic for all 367 cities, we have the following findings:

**Figure 5:**
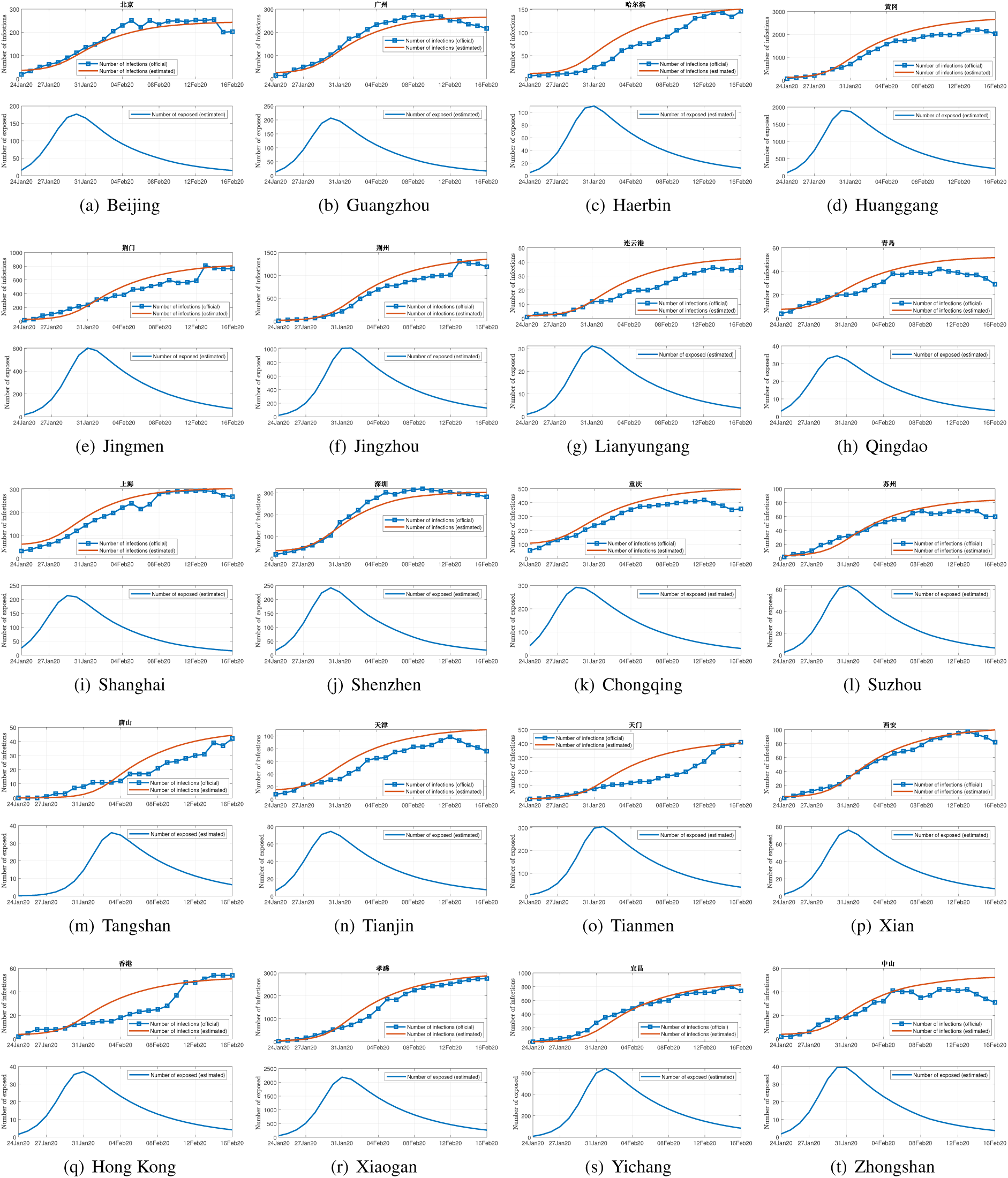
Official number of infected individuals and estimated number of infected individuals in 20 selected cities in China (upper), and estimated number of exposed individuals (lower).

**Figure 6:**
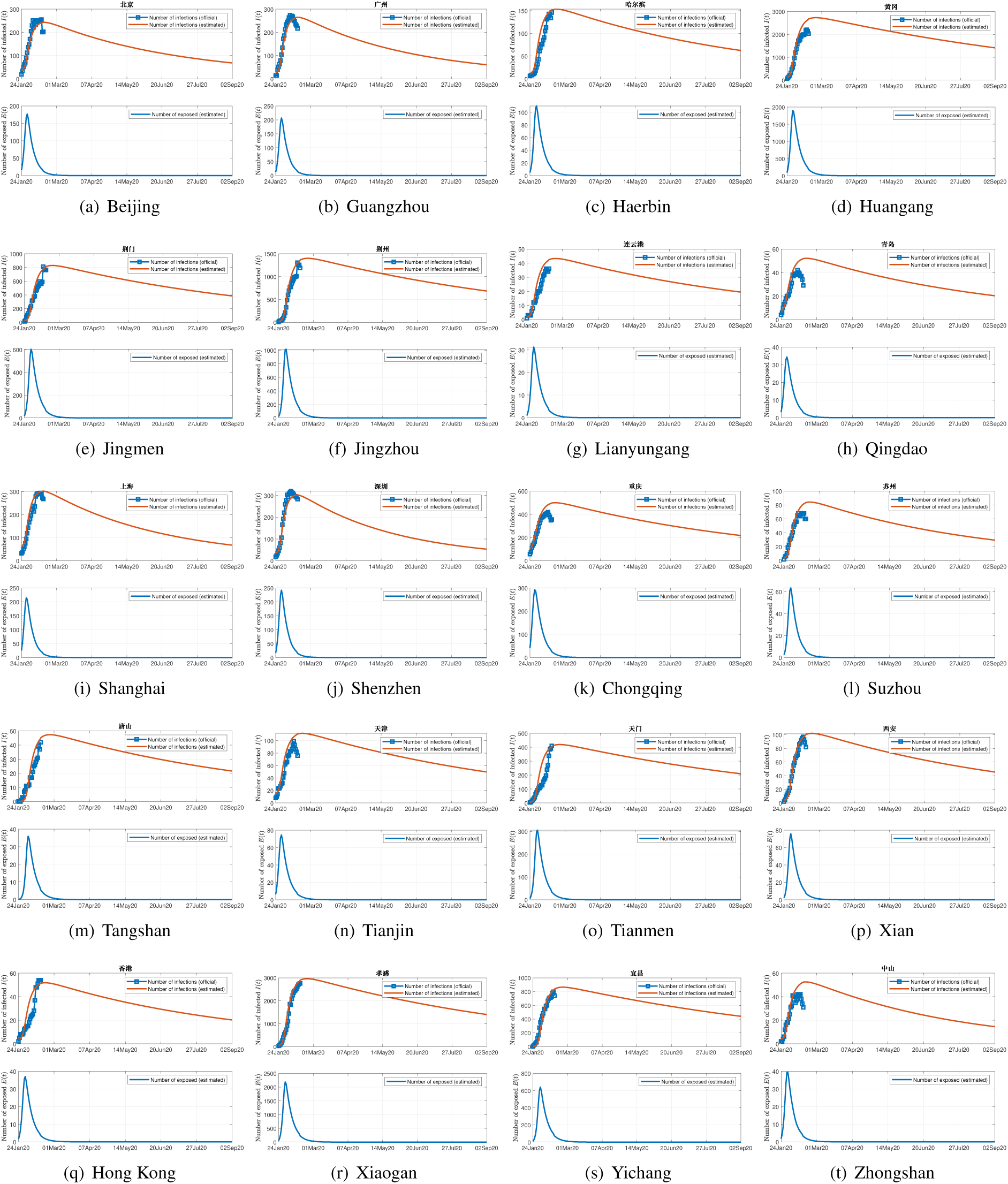
Prediction of the number of infected (upper) and exposed individuals (lower) in 20 selected cites in China for the next 200 days Chongqing for the next 200 days.

- For most cities in China, the infection numbers will peak between mid February to early March 2020, as shown in Figure 7(a).
- The peak number of infected individuals will be between 1,000 to 5,000 for cities in Hubei, and that outside Hubei will be below 500, as shown in Figure 7(b).
- At the end, about 0.8%, less than 0.1% and less than 0.01% of the population will get infected in Wuhan, Hubei Province and the rest of China, respectively, as graphically presented in Figure 7(c).

Opinions diverge in the social media on the estimated extent of the outbreak of the new coronavirus disease (COVID-19). While there are pure speculations, there are also predictions based on rigorous study of the spreading dynamics. Different models used for prediction and different assumptions made regarding the transmission process would lead to different results and quite diverged conclusions. For instance, an AI-powered simulation run had forecasted 2.5 billion people to be infected in 45 days [13]. Academics in Hong Kong expected 1.4 million eventually infected in the city of 7.5 million people. Our results, however, do not seem to agree with such forecasts. In fact, our results are expected to be conservative, under normal circumstances, in the sense that the severity and duration of the epidemic could be over-estimated. This is because stringent measures have been taken to limit travel and hence to curb the spreading of the virus since late January 2020. The infection rates should be progressively reduced, with the effect of reducing the number of infected individuals as well as mitigating of the peak times. Furthermore, climate factors may also play a role in the epidemic spreading. As temperature generally rises from February to May, the infection peaks would unlikely occur later than the predicted times as the new coronavirus epidemic is generally known to subside in warm weather. Based on our findings and assuming no drastic change in infection rates (e.g., schools and workplaces remain closed and intercity travel limited), we can conclude, with optimism, that the COVID-19 epidemic would end around April 2020.

**Figure 7:**
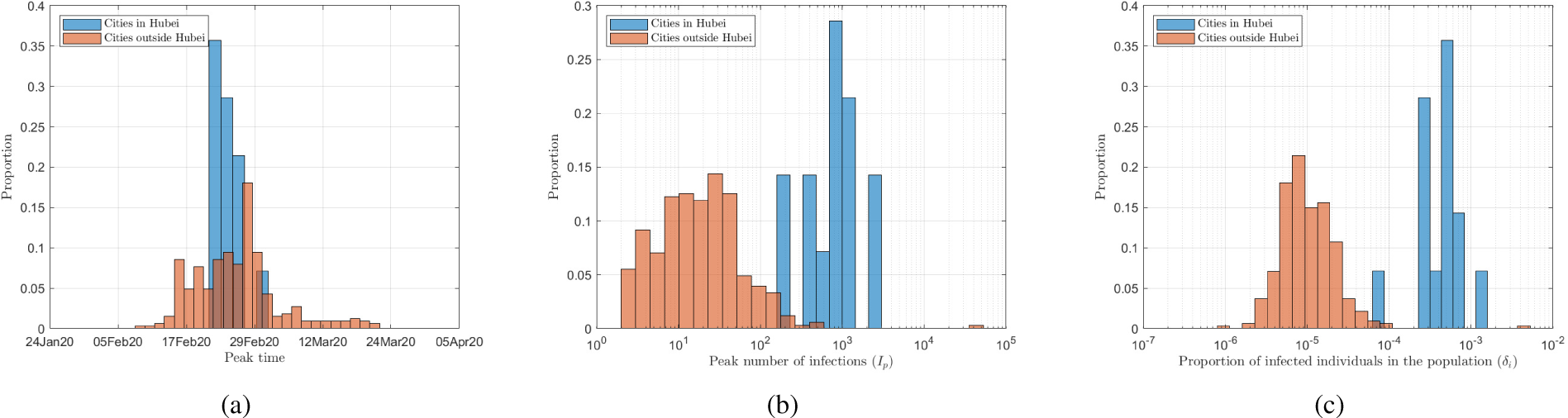
(a) Distribution of peak time; (b) distribution of peak number of infections; (c) distribution of proportion of the population eventually infected in a city.

## 5. Conclusion

We employ human migration data to provide information on intercity travel that is crucial to the transmission of the new coronavirus disease from its epicenter Wuhan to other parts of China. Our model for the disease spreading is essentially the classic SEIR model, with intercity travel data supplying the essential information about the number of infected, exposed and recovered individuals moving between different cities. All parameters of the model, including infection rates, recovery rates, and eventual percentage of infected population for 367 cities in China, are identified by fitting the official data with the model using a constrained nonlinear programming procedure. Using these parameters, estimates of the number of exposed individuals in 367 cities as well as projections into the next 200 days are also found. Our model, however, does not consider the contact network topology that would be necessary if details of the transmission process, such as superspreading events, are to be captured. Nonetheless, our model provides a very reasonable estimation of the propagation of average numbers of exposed, infected and recovered individuals, despite missing details of fluctuation (e.g., sudden surge due to a superspreading event). The main conclusion of our study is that the COVID-19 epidemic spreading will peak between mid February to early March 2020, with about 0.8%, less than 0.1% and less than 0.01% of the population eventually infected in Wuhan, Hubei Province and the rest of China, respectively.

## Data Availability

The human migration data are obtained from Baidu Migration (link: http://qianxi.baidu.com), and the COVID-19 infection data for China are provided by National Health Commission of China and available from https://ncov.dxy.cn/ncovh5/view/pneumonia.

https://ncov.dxy.cn/ncovh5/view/pneumonia

## Acknowledgment

This work was supported by National Science Foundation of China Project 61703355 and Guangdong Youth University Innovative Talents Project 2016KQNCX223, and City University of Hong Kong under Special Fund 9380114 provided by VP(RT).

